# *I couldn’t confirm they were alone because they were over the phone*: A qualitative study of safeguarding during remote sexual and reproductive health consultations in England and Wales

**DOI:** 10.64898/2026.02.03.26345229

**Authors:** Charlotte Spurway, Tom Witney, Helen Munro, Jo Josh, Melvina Woode Owusu, Jo Gibbs, Iestyn Williams, Danielle Solomon, Andrew Copas, Jonathan DC Ross, Louise Jackson, Fiona Burns

## Abstract

Remote consultations, including telephone, video, text or web-based consultations, are now common in sexual and reproductive health services (SRHS) in England and Wales, offering convenience and efficiency but raising concerns about safeguarding service users compared to in-person care. Ensuring protection and support for individuals remain central to SRHS. While guidance exists, evidence on how safeguarding is enacted remotely is limited. This study explores how safeguarding is managed in remote SRHS, examining associated challenges, benefits, and perceptions of acceptability. This qualitative study, part of the CONNECT study (NIHR153151), explored safeguarding in remote delivery of SRHS across three diverse case study areas in England and Wales. Semi-structured interviews were conducted with service users and providers (March 2024-January 2025). Data were thematically analysed using inductive and deductive approaches. Eighty-nine interviews were conducted with 54 service users/potential service users and 35 professional stakeholders across three study sites. Four key themes were identified: (1) challenges of delivering remote safeguarding in practice, (2) importance of a safe space for the patient, (3) one size approach does not fit all, and (4) strategies to support remote safeguarding. Participants described difficulties identifying risks without visual cues, variable comfort with disclosure, and privacy concerns. Providers used adaptive, collaborative approaches to build trust, ensure safety, and tailor safeguarding to individual circumstances. Both service users and providers recognised the effectiveness of remote safeguarding largely depended on clinician judgement, structured questioning, and sensitive communication. While remote consultations offered flexibility, remote safeguarding relied on service users having access to safe, private space. People with language barriers or those less able to create a safe space at home, such as those experiencing housing instability or coercive control may also have greater safeguarding needs. Ensuring access to in-person options, clear safeguarding protocols, and appropriate clinician training is essential to mitigate these challenges.

**Author summary:** During the COVID-19 pandemic, sexual and reproductive health services used more remote appointments, such as phone, video, or online consultations. These appointments can make it easier for some people to get care. But there are concerns about whether staff can spot safeguarding issues, such as people who may be at risk of harm, without seeing them in person. In this study, we spoke with people who use sexual and reproductive health services, people who had never used these services before, and staff who work in these settings. People described challenges with remote appointments, including not being able to see body language, feeling unsure about talking about sensitive issues, and worries about privacy at home. Staff described changing how they worked by asking careful questions, building trust, and making safeguarding fit each person’s situation. Both service users and staff felt that keeping people safe during remote appointments depends on clear communication and professional judgement. It is harder when people do not have a private space, face language difficulties, or live in unsafe housing or controlling relationships. Our findings show the importance of keeping in-person appointments available and making sure staff have the right training and guidance.

## Introduction

Remote consultations, conducted via telephone, video, text or web-based consultations, are now routine in Sexual and Reproductive Health Services (SRHS) in England and Wales [1]. This form of consultation has been shown to provide benefits such as convenience, privacy, and anonymity [1, 2] and the potential to address capacity and resource constraints and increase efficiency [3]. However, evidence suggests remote consultations may miss important information compared to in-person appointments, such as the ability to perform physical examinations and to identify safeguarding concerns and confidentiality issues [4-6].

Safeguarding is a fundamental part of the UK health and social care system, with service providers responsible to protect individuals from harm, abuse and neglect [7]. It encompasses activities designed to protect individuals, guided by principles of empowerment, protection from harm, proportionality through the least intrusive response, and accountability [8]. While all patients may be at risk, the NHS identifies ‘vulnerable groups’, who require particular attention due to factors such as age, disability, cognitive or mental health impairment, dependence on others, social displacement, experiences of abuse or exploitation, or structural marginalisation [9].

Within SRHS, safeguarding is a key aspect of delivering care by ensuring safety, well-being, and access to appropriate support. This includes routine screening of people under-18 for child sexual abuse [10], as well as assessing whether they are competent to make decisions about medical treatment, such as abortion care [11]. In addition, adults are routinely screened for intimate partner violence. During the COVID-19 pandemic, UK abortion provision shifted to remote consultations and at-home use of abortion medication, a change introduced in 2020 and made permanent in England and Wales in 2022 [12]. However, some groups have raised concerns about remote consultations for abortion on safeguarding grounds, arguing that remote provision may limit healthcare professionals’ ability to identify abuse or neglect [12].

Existing research indicates that in online SRHS, safeguarding referrals for under-18s are most often triggered by drug or alcohol use, partner age imbalances, or involvement with social or mental health services [2]. However, findings have been mixed regarding how effectively safeguarding operates in remote SRHS contexts. Much of the existing SRHS literature focuses on online asynchronous interactions, such as response to online assessment questions used for postal self-sampling. Online sexual history taking may encourage individuals, particularly young people, to disclose personal or embarrassing information, suggesting that remote services could facilitate reporting of safeguarding-related concerns [10].

The COVID-19 pandemic highlighted both the potential strengths and limitations of remote safeguarding with concerns that certain safeguarding issues may have remained undisclosed during lockdown periods [13]. Research from primary care indicates that remote consultations can complicate history-taking and dialogue, potentially exacerbating inequities for vulnerable groups, particularly for those with intersecting factors, such as older adults with limited English and digital literacy or individuals facing poverty alongside multiple health conditions [14]. Much of the literature has focused on individual’s under-18 reflecting the legal framework governing safeguarding in SRHS, but there is also limited research exploring remote safeguarding from the perspective of vulnerable adults.

Several SRHS organisations provide guidance to support safe remote safeguarding practices. The Faculty of Sexual and Reproductive Healthcare ((FSRH) (now College of Sexual and reproductive Healthcare, CoSRH)) and the British Association for Sexual Health and HIV (BASHH) [15] emphasise that thorough risk assessments, combined with clear escalation pathways are important to address safeguarding concerns [15]. The British HIV Association (BHIVA) [16] highlights the importance of confirming patient identity, safety to talk and secure location at the start of consultations, while the Royal College of Obstetricians and Gynaecologists (RCOG) [17] stresses the need to remain alert to verbal cues and conduct robust assessments, particularly for vulnerable groups such as under-18s, those with disabilities, or individuals misusing substances. The CoSRH and BASHH [18] emphasise staff training to recognise risks and ensure remote triage protocols include guidance for timely in-person referrals or safeguarding actions. Despite this guidance, understanding how safeguarding is navigated in practice from both professional stakeholders and service user perspectives. The aim of this study was to explore safeguarding practices and the challenges, benefits and acceptability of these processes in remote service delivery.

## Materials and methods

### Study design

This study forms part of the CONNECT study (NIHR153151), which explored the impact of remote consultations on health inequalities in SRHS. We conducted semi-structured interviews in three case study areas (CSAs) in England and Wales. A section of the interview focused on safeguarding and the associated findings are discussed in this paper. The three CSAs were selected to capture sociodemographic diversity and to explore contrasts between rural and urban settings. Table 1 summarises the appointment types across the three CSAs, highlighting each area’s approach to in-person and remote consultations. Interviews lasted 60 minutes and were conducted between March 2024-January 2025.

**Table 1.** Overview of appointment and consultation options by service.

|  | CSA A | CSA B | CSA C |
| --- | --- | --- | --- |
| Walk-ins | Under 25s only | þ | X |
| Booked appointments | þ | þ | þ |
| Online postal self-sampling | X | þ<br>(in-house provision) | þ<br>(external provider) |
| F2F consultation | þ | þ | þ |
| Remote consultation | Limited telephone appointments | þ | þ |

F2F = In-person face to face consultation, X = not offered/not available by the service, þ = available at the service.

For the purposes of this study, remote consultations are defined as synchronous interactions between patients and clinicians that do not require in-person contact. These include telephone or video consultations, as well as secure two-way written communication (e.g. SMS, email, or online messaging platforms). Asynchronous consultations delivered through online postal self-sampling were excluded, as these have been examined previously [19].

### Sampling and recruitment

The study employed a maximum variation sampling technique and included service users from a diverse range of backgrounds, including ethnicity, age, gender sexual orientation, socioeconomic status, and disability [20]. Potential service users were included to capture diversity of experience and reasons for non-use. A similarly wide range of professional stakeholders, including service providers, managers and commissioners were included, representing different clinical and non-clinical roles, levels of experience and perspectives.

Our recruitment target was 15-20 service users/potential service users and 10-15 professional stakeholders from each of the three CSAs, with recruitment continuing until data saturation was achieved. Participants were recruited through multiple methods including: posters displayed in SRHS clinics, in-person approaches by interviewers and local research teams in clinic settings, outreach via established networks, email invitations, text messages to recent service users, and the CONNECT study website. All recruitment materials were provided in English and Welsh.

Service users/potential service users were eligible if they were aged 16 years or older and resided within one of the CSAs. Participants also needed to have the capacity to provide informed consent and to speak and read English, as interviews could not be conducted in other languages. Professional stakeholders were eligible if they were currently involved in delivering SRHS within the three CSAs. Eligible professional stakeholders included: consultants, nurses, health advisers, GPs, other physicians or clinicians, counsellors, managers, commissioners or those involved in SRHS delivery.

### Data collection

Participants were invited to take part in an interview after reviewing the participant information leaflet and providing informed consent. Participants also completed a screening questionnaire to capture socio-demographic information to support sample diversity. Most of the questions were optional, and participants could select ‘prefer not to say’ for any question.

Interviews were conducted by an experienced qualitative researcher (CS or TW). To encourage participation, interviews were conducted in participant’s preferred format, either in person at an SRHS, on university premises, or remotely via telephone, Microsoft Teams, or Zoom. All interviews were audio-recorded using an encrypted Dictaphone or built-in recording functions in Microsoft Teams or Zoom.

Interviews with service users/potential service users followed a topic guide exploring experiences of SRHS, including access, the use of remote versus in-person consultations, and barriers to care. The topic guide for service users/potential service users was piloted with the CONNECT Voices PPI panel. Participants who had never used the service were asked about their reasons for non-use, including lack of need, barriers to access, or alternative service use, and what might improve access. For professional stakeholders, interviews focused on perspectives of SRHS consultations in both remote and in-person formats. The findings from this component of the study are reported and discussed in a separate paper [21].

The safeguarding component examined participants’ experiences of being asked or asking about domestic and/or sexual violence during remote consultations, as well as their comfort with such questions. For professional stakeholders, this included discussing experiences of asking safeguarding questions, while those without direct consultation experience such as other stakeholders and commissioners were asked about their perceptions, knowledge, and thoughts.

### Data analysis

Audio recordings were transcribed verbatim by a university-approved transcription company and pseudonymised prior to analysis. A thematic analysis was conducted in two stages. First, as part of a broader qualitative study, transcripts were coded by CS and TW using combined inductive and deductive approaches. During this initial coding phase, data relating to safeguarding were labelled under a broad safeguarding code and did not form the main focus of the primary analysis. Second, for the purposes of the present paper, all data coded as relating to safeguarding were re-examined and analysed in greater depth. This subset of data was re-coded inductively capturing participants’ safeguarding-related experiences and perspectives. These codes were discussed within the wider research team and iteratively refined and grouped into broader analytic themes, which form the basis of the findings presented here. NVivo v.14 (Lumivero, Burlington, MA) was used for data management and analysis.

### Ethical considerations

The study received ethical approval from the NHS Research Ethics Committee (REC: 23/NS/0128). Participants gave informed consent before participation, either in person using paper forms or electronically via REDCap (Research Electronic Data Capture; Vanderbilt University, Nashville, TN), in accordance with the approved ethical protocol. A £20 shopping voucher was provided to service users/potential service users in acknowledgment of their contribution.

## Results

In total, we carried out 89 interviews with 54 service users/potential service users and 35 professional stakeholders across the three CSAs. Tables 2 and 3 display the characteristics of participants who took part in the study.

**Table 2.** Service user/potential service user characteristics.

|  |  | CSA A | CSA B | CSA C | Overall |
| --- | --- | --- | --- | --- | --- |
| Age | 16-20 | 2 | 3 | 3 | 8 |
|  | 21-24 | 2 | 4 | 2 | 8 |
|  | 25-34 | 10 | 5 | 8 | 23 |
|  | 35-44 | 1 | 2 | 3 | 6 |
|  | 45+ | 6 | 3 | 0 | 9 |
| Gender | Cisman | 8 | 4 | 4 | 16 |
|  | Ciswoman | 9 | 10 | 8 | 27 |
|  | Transman | 1 | 0 | 1 | 2 |
|  | Transwoman | 2 | 0 | 1 | 3 |
|  | Non-binary person | 1 | 2 | 1 | 4 |
|  | Prefer not to say | 0 | 1 | 1 | 2 |
| Ethnicity | Asian or Asian British | 2 | 4 | 0 | 6 |
|  | Black/African/Caribbean/Black British | 3 | 4 | 0 | 7 |
|  | Mixed/multiple ethnic groups/other | 3 | 1 | 2 | 6 |
|  | White | 13 | 8 | 14 | 35 |
|  | Prefer not to say | 0 | 0 | 0 | 0 |
| Sexuality | Heterosexual or straight | 9 | 6 | 9 | 24 |
|  | Gay or Lesbian | 6 | 7 | 2 | 15 |
|  | Bisexual | 6 | 2 | 3 | 11 |
|  | Other | 0 | 2 | 2 | 4 |
|  | Prefer not to say | 0 | 0 | 0 | 0 |
| Disability<sup>a</sup> | Yes | 7 | 8 | 3 | 18 |
|  | No | 14 | 9 | 12 | 35 |
|  | Prefer not to say | 0 | 0 | 1 | 1 |
| Education | Secondary or earlier | 1 | 0 | 1 | 2 |
|  | Further education | 3 | 4 | 4 | 11 |
|  | University/postgraduate | 17 | 13 | 11 | 41 |
|  | Prefer not to say | 0 | 0 | 0 | 0 |
| Ever used SRHS services | Yes | 20 | 12 | 11 | 43 |
|  | No | 1 | 4 | 5 | 10 |
|  | Prefer not to say | 0 | 1 | 0 | 1 |
| Prior diagnosis of an STI | Yes | 10 | 3 | 2 | 15 |
|  | No | 11 | 14 | 13 | 38 |
|  | Prefer not to say | 0 | 0 | 1 | 1 |
| **Overall** |  | **21** | **17** | **16** | **54** |

<sup>a</sup>Participants response to the question “Do you have any physical or mental health conditions or illnesses lasting or expected to last 12 months or more?”

**Table 3.**
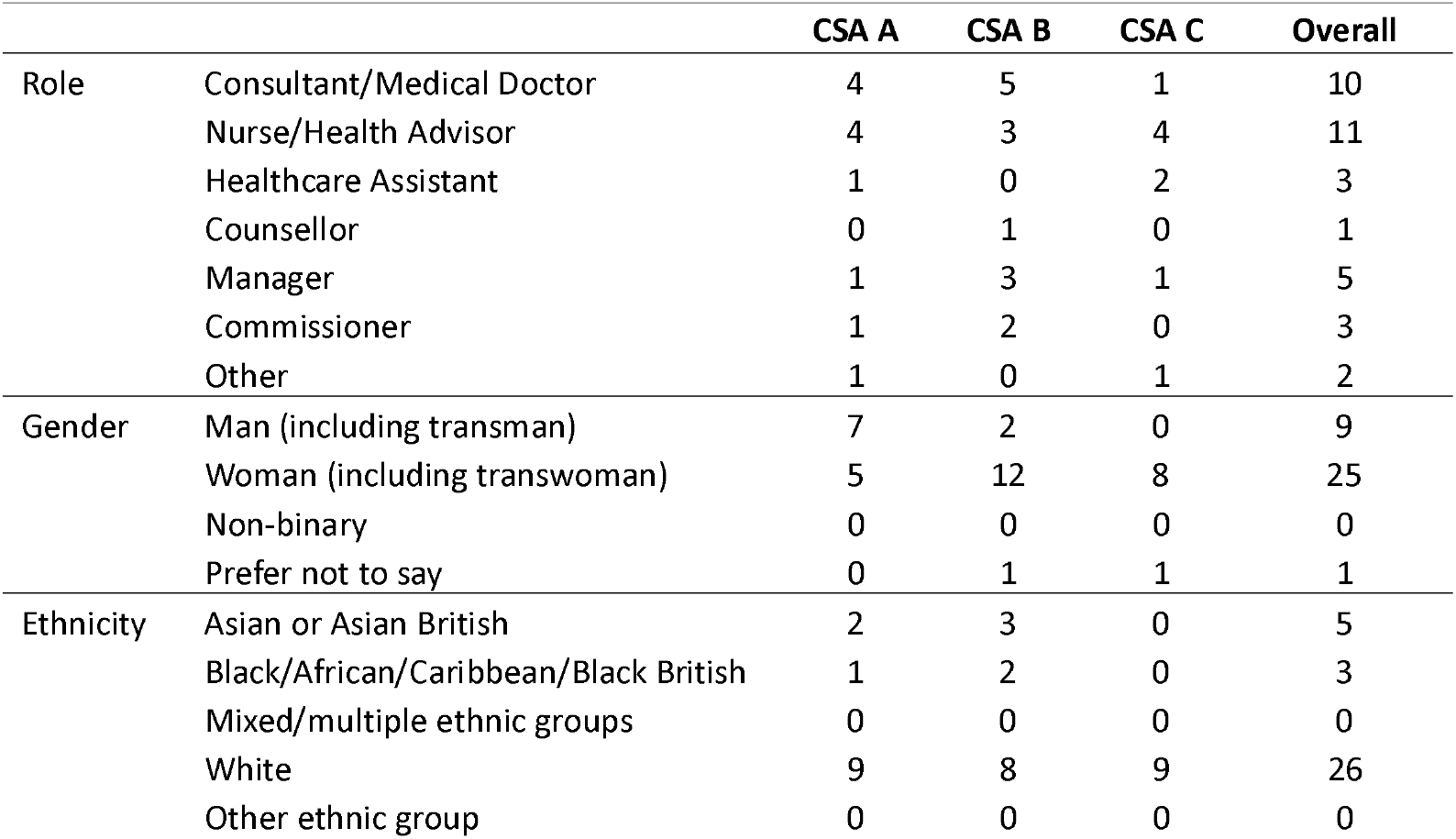
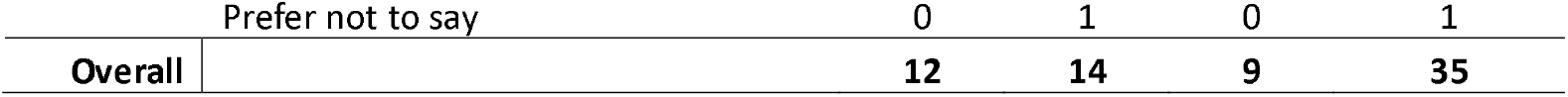
Professional stakeholder characteristics.

|  |  | CSA A | CSA B | CSA C | Overall |
| --- | --- | --- | --- | --- | --- |
| Role | Consultant/Medical Doctor | 4 | 5 | 1 | 10 |
|  | Nurse/Health Advisor | 4 | 3 | 4 | 11 |
|  | Healthcare Assistant | 1 | 0 | 2 | 3 |
|  | Counsellor | 0 | 1 | 0 | 1 |
|  | Manager | 1 | 3 | 1 | 5 |
|  | Commissioner | 1 | 2 | 0 | 3 |
|  | Other | 1 | 0 | 1 | 2 |
| Gender | Man (including transman) | 7 | 2 | 0 | 9 |
|  | Woman (including transwoman) | 5 | 12 | 8 | 25 |
|  | Non-binary | 0 | 0 | 0 | 0 |
|  | Prefer not to say | 0 | 1 | 1 | 1 |
| Ethnicity | Asian or Asian British | 2 | 3 | 0 | 5 |
|  | Black/African/Caribbean/Black British | 1 | 2 | 0 | 3 |
|  | Mixed/multiple ethnic groups | 0 | 0 | 0 | 0 |
|  | White | 9 | 8 | 9 | 26 |
|  | Other ethnic group | 0 | 0 | 0 | 0 |

|  | Prefer not to say | 0 | 1 | 0 | 1 |
| --- | --- | --- | --- | --- | --- |
| **Overall** |  | **12** | **14** | **9** | **35** |

Four key themes were identified: (1) Challenges of remote safeguarding in practice; (2) Importance of a safe space; (3) One size does not fit all; and (4) Strategies to support remote safeguarding.

### Challenges of remote safeguarding in practice

Safeguarding assessments were routinely included in both in-person and remote consultations, often at the end of sexual history taking. While experiences varied by modality, some providers felt effectiveness depended more on clinicians’ professional curiosity than the consultation format. As one participant explained:

> *‘I don’t think virtual working has a detrimental effect on safeguarding whatsoever. I think our staff are really good at using their professional curiosity whether it’s in a virtual consultation or a face-to-face consultation to get to the bottom of what’s going on*.*’ (HCP07, Nurse/Health Advisor)*

However, some clinicians reported more avoidance of safeguarding related questions during remote consultations than when in-person. Over the phone, it was easier to skip questions if the conversation felt rushed, uncomfortable, or if the clinician was unsure how to frame them.

> ‘*But I think sometimes it’s a bit of a get out clause from asking what can be quite difficult questions and it’s, oh well I couldn’t confirm they were alone because they were over the phone, you know*.’ (HCP03, Nurse/health advisor)

Remote consultations also raised concerns about incomplete context and reduced privacy. Some worried that remote consultations could miss important non-verbal cues, such as body language, facial expressions, and tone of voice, that help identify underlying concerns. One clinician highlighted that remote consultations could limit opportunities to ask safeguarding questions, particularly when there was uncertainty about a patient’s privacy. Considering the potential for missing information during remote consultations, participants emphasised that disclosure is voluntary for those with capacity, and further intervention is not always possible without explicit consent.

> *‘Ultimately, if you’ve done everything you can do to keep them safe, if you’ve done the referrals and made sure that everything is in place for them, then*… *we’ve just got to accept that there’s nothing else that you can do at that point*.*’* (HCP21, Nurse/health advisor)

Service users/potential service users expressed mixed views. Some were sceptical about disclosing concerns over the phone or online, worrying that these formats lacked sufficient support. As one participant explained, the absence of in-person support could leave users feeling vulnerable:

> ‘*You could answer those questions and be a gibbering wreck at the end of it and then you hang up the phone…Whereas I think if you’re doing it in a room with a bunch of professionals and it did trigger a meltdown there’s people there to comfort you and support you*.’
>
> (SU04, cisman, age: 45+)

Across both groups, trust was central to disclosure. One-off questions were seen as insufficient, and disclosure depended on timing, rapport, and readiness. Remote modalities, especially short calls, were described as less conducive to building the trust and relationship needed for people to feel able to disclose.

> *‘I keep asking about domestic violence on a telephone appointment they say, ‘No*.*’ and they said, ‘Well you ask me every single time The day I see you face-to-face I’ll tell you all about it*.*’ and it took someone two years to come back and tell me*.*’* (HCP32, Consultant/medical doctor)

### Importance of a safe space

Participants emphasised that in-person consultations offered privacy, confidentiality, and security that supported trust-building and open communication. Being in a clinical environment, away from home, made disclosure feel safer, especially for those experiencing control, coercion, or abuse.

> ‘*I think possibly I would feel more comfortable in a face-to-face appointment. Also I mean having distance, being in a separate location*.’ (SU39, ciswoman, age: 25-34)

Clinicians similarly highlighted that in-person settings allowed them to create and manage a safe environment, for example by ensuring privacy or asking a partner to wait outside. This physical separation was essential for maintaining confidentiality and facilitating safeguarding conversations. In contrast, remote consultations made it harder to guarantee privacy or control the setting, meaning providers could not always be sure a patient was alone or able to speak freely.

> ‘*We don’t know who’s listening in on the virtual consultation, sometimes there’s someone visibly in the background and you can’t really do anything about that you know? You can’t control that like you can in the clinic*.’ (HCP36, Consultant/medical doctor)

This concern was further highlighted by another clinician highlighted concerns when others were present during remote consultations, leading them to rely on ‘gut feeling’ to prompt further enquiry.

> ‘*And also, sometimes I find somebody else might be in the room with them, and you may not know who that person is…Or they’ll just say, ‘oh, it’s my friend’. But then that friend is sometimes a little bit older than them, and- You know, yeah. And then you have this gut feeling*.’ (HCP04, Nurse/health advisor)

Service user/potential service user perspectives varied. While many valued the security and structure of clinic settings, others felt more comfortable disclosing sensitive information remotely. For these individuals, physical distance reduced embarrassment or intimidation, creating a sense of psychological safety and control that allowed them to open up at their own pace in a familiar environment.

> ‘*I think a remote consultation would, at least for me, make it more able and less like distressing for me to like get into detail about this kind of stuff because I think it can feel quite overwhelming if it’s like a person in front of me asking me*.’ (SU65, ciswoman, age: 21-24)

### One size does not fit all

The interviews found differences in how adults and under-18s are managed across all the CSAs. For under-18s, in-person appointments were prioritised regardless of reason for consultation, underpinned by legal safeguarding responsibilities. As one clinician explained:

> ‘*We do try and attempt numerous times, but there’s only so much you can do. They’re an adult at the end of the day, so there’s limits to what we can do, whereas with under-18s we can do a safeguarding referral without their permission*.’ (HCP19, Consultant/medical doctor)

In cases involving under-18s, any safeguarding concerns raised during a remote consultation were more likely to prompt a follow-up in-person appointment. This reflects a lower threshold for in-person assessment for younger patients, whose needs and risks require closer observation and intervention.

> ‘*I think with young people, if a young person made a disclosure that was concerning around safeguarding over the phone, then we would probably aim to get them to come in for a face-to-face appointment with our health advisors*…’ (HCP29, Consultant/medical doctor)

Remote consultations can offer discreet access to services for all users. Participants noted that for someone experiencing domestic violence, remote appointments might allow them to seek help when alone or at lower risk. However, they also imagined that in severe cases, constant monitoring could make private access impossible.

> ‘*Right now, on one side that person might be able to just organise the call with you at a time when the risk is lowest or when they are on their own. I mean it really depends on how severe the situation is*’ (SU24, cisman, age: 45+)

Service users reflected that comfort with remote consultations depends on personal risk and past experiences. Those without experiences of domestic abuse, sexual violence, exploitation, sex work or substance use generally felt confident answering safeguarding questions remotely and perceived little difference between formats.

> ‘*For me, it didn’t really make a difference, because I haven’t experienced domestic abuse or anything. I didn’t think anything of it. It’s the same for me, whether it’s face-to-face or over the phone. Those kinds of questions. I just think it might be different if you were actually affected by that*.’(SU22, ciswoman, age: 35-44)

Participants noted that preferences for mode of consultation varied. From a practical perspective, remote consultations could facilitate disclosure for those facing barriers to attending in person, such as childcare, mobility, or mental health challenges.

> ‘*Although we talk about the bad parts of missing vulnerabilities and things if we do remote consultations, it can also be helpful. So if you have young children or you’re in a coercive relationship, it might be beneficial’*. (HCP31, Consultant/medical doctor)

### Strategies to support remote safeguarding

Professional stakeholders described using various strategies to identify and respond to safeguarding concerns remotely. They cautiously introduced sensitive topics after assessing privacy, safety, and the individual’s environment, attended to verbal cues such as tone, background noise, and hesitations, and adjusted their approach accordingly. Building rapport was seen as important, with some delaying safeguarding discussions until trust was established.

> ‘*I would normally wait and build a rapport with the patient*… *Then I would normally give people a warning and say, ‘I’m going to be asking you some questions about domestic violence and sexual abuse are you still somewhere safe and private to speak where you can answer freely?‘*’ (HCP31, Consultant/medical doctor)

Participants supported a collaborative, patient-centred approach, discussing concerns openly, explaining the limits of confidentiality, and co-creating safety plans where possible and working carefully to minimise harm through joint decision-making. Flexibility and creativity were also seen as important. Alternative communication methods such as text messages, letters, or follow-up calls were suggested. These were seen as helping to maintain contact, ensure information and support are accessible, and address barriers to safe disclosure.

> *‘I suppose like if someone isn’t the best with their phone, you could send them a letter if you were doing it remotely and then they’ll have the information and any support available. There’s ways around it. You just have to think a little bit differently*.’ (HCP21, Nurse/health advisor)

In response to safeguarding concerns, some clinicians arranged in-person appointments under alternative pretences, allowing discreet in-clinic conversations.

> ‘*The whole thing was engineered round a smear. To get the woman in to be able to discuss it. She didn’t need to smear but it was broached in that that would have been very difficult to do. Have access to… he wouldn’t give access to a phone*.’ (HCP08, Nurse/health advisor)

One participant suggested that remote safeguarding could be improved by offering greater flexibility and availability, allowing individuals to engage at times when they feel safe to talk.

> ‘*I think it’s just a matter of scheduling and being available to having more availabilities around when the person might be safe to talk*.’ (SU81, ‘prefer not to say’, age: 16-20)

## Discussion

This study identified four key themes related to safeguarding in remotely delivered SRHS across three CSAs. Both service users/potential service users and professional stakeholders recognised the complexities of safeguarding in remote contexts, alongside some potential benefits. Many participants did not express strong preferences for remote or in-person consultations, rather the perceived effectiveness of remote safeguarding depended on clinicians’ professional curiosity and responsiveness. However, it is important to note that preferences are not always entirely free choices, particularly for young people or those experiencing abuse, as they may be shaped by coercion, trauma, or practical constraints. Making space for these influences is important for designing services that are responsive to patient needs. This corresponds with existing evidence that professional judgement and training remain central to effective safeguarding across all modalities, although applying this judgement may be more challenging remotely [22]. Remote delivery can also introduce barriers that affect clinicians’ confidence and consistency when raising safeguarding concerns, especially during sensitive or time-pressured conversations. Strategies such as structured questioning, sensitive communication, and attentive listening can support disclosure remotely [23].

Despite these strategies, participants also described obstacles that impact safeguarding in practice. These include failing to identify relevant information and the lack of non-verbal cues. Professional stakeholders in this study reflected that remote consultations can make it harder to be completely confident that service users are able to disclose safeguarding concerns fully, echoing prior research suggesting that risk assessments conducted remotely are less reliable than those conducted in-person [24, 25]. This has important implications for SRHS that do not offer in-person appointments, for example, remote abortion care, where potential risks may be greater. However, providers noted that safeguarding in SRHS is an ongoing process and that, when needed, further assessment can be deferred to a later in-person appointment, after an initial remote consultation.

For adults, disclosure remains dependent on their willingness and ability to share information; if a patient who has capacity chooses not to disclose safeguarding-related matters, SRHS are limited in the actions they can take. The consultation environment and patients’ sense of privacy and confidence in the service can influence their ability to share sensitive information. For example, limited private space for remote consultations, such as for people in shared housing, multi-generational households or in coercive relationships, can restrict confidentiality, as previous studies have shown [24, 26, 27]. One study found that predictability, such as fixed appointment times, is important for remote consultations as it allows patients to prepare and create a safe space [28].

Service users also expressed concerns about how disclosures would be handled remotely, highlighting the need for clear information about processes and expected outcomes. In-person consultations were associated with a sense of privacy and security, with the clinical setting perceived as a controlled environment that facilitates open communication. This aligns with previous work identifying consulting rooms as safe spaces for discussing sensitive issues [22]. Remote consultations, however, can offer distinct benefits, particularly in providing patients with greater control over the interaction. Service users suggested it would allow them to choose when to engage, have greater control over the pace of the conversation and feel less constrained by an unfamiliar clinical environment. This flexibility may support disclosure, especially for individuals discussing trauma, abuse, or coercive relationships [23].

Age-specific considerations were also evident. Legal frameworks mean that safeguarding for under-18s allows more proactive intervention once concerns are identified, whereas safeguarding adults generally relies on voluntary disclosure and their consent to any intervention. This affects decisions around consultation type, follow-up, and the approach to risk assessment. Remote consultations can offer safeguarding benefits for vulnerable groups defined by the NHS, such as young people, individuals with physical, sensory, or mental impairments, and those with learning disabilities [9], by improving access when in-person visits are difficult. However, challenges remain in supporting disclosure and assessing risk effectively [22, 24]. If remote safeguarding is the only option, individuals may struggle to disclose information due to limited access to the service or lack of private, safe space. Offering a choice of consultation type is therefore essential, with in-person services important when privacy is needed.

Professional stakeholders described a range of strategies to support remote safeguarding, including structured questioning, ensuring patient privacy, and planning for a follow-up review or in-person consultation. While formal frameworks were not widely discussed in interviews, services have safeguarding protocols to guide clinicians. Many of these strategies align with guidance from professional bodies such as BASHH, BHIVA, and CoSRH. These recommendations emphasise thorough risk assessment, ensuring confidentiality, and having clear procedures for escalation, suggesting that provider practices reflect established professional standards. The broader findings of this study also recognise that not all patients have ready access to the digital technologies needed for remote consultations [21]. Ensuring that patients have the option for in-person contact where needed, providing clear information about safeguarding processes, and tailoring approaches to individual patient circumstances are essential to delivering safe and effective care [25]. Building on these findings, future research could examine how different remote consultation modalities impact actual safeguarding outcomes and explore which approaches best support disclosure across diverse patient groups and circumstances. Understanding these nuances can inform service design, ensuring that remote and in-person consultations complement each other and that safeguarding processes remain responsive to patient needs.

### Strengths and limitations

The strengths of this study include the inclusion of service users, non-users of SRHS, commissioners, and providers from three diverse CSAs, allowing exploration of remote safeguarding from multiple perspectives. Additionally, by recruiting a diverse sample of participants across these CSAs, the study was able to capture the experiences of individuals from marginalised communities in relation to safeguarding practices.

This study is part of a wider investigation into the impact of remote consultations in SRHS on health inequalities. It is important to note that service users generally had limited experience of disclosing information that triggered safeguarding concern as we did not specifically sample individuals with such experience. Many participants reported that they had never raised issues related to safeguarding. In some cases, service users may have been invited to in-person consultations due to safeguarding concerns but were unaware that this was the reason for the invitation. The study also included a limited sample of under-18s, as participants had to be aged 16 or older to take part, and younger individuals may have different experiences of safeguarding within SRHS. In addition, people with disabilities such as hearing or sight loss may have found virtual consultations particularly challenging, and the study’s requirement for sufficient English-language comprehension may mean that the findings underrepresent key equity concerns.

## Conclusion

Safeguarding within remote SRHS consultations was shown to have benefits, but its effectiveness depends on clinician judgement, a structured approach and clear communication. The ability to exercise such judgement may be limited by factors including training, time available, and the working environment. Remote consultations offer benefits such as flexibility, patient control, and improved access for some vulnerable populations, but challenges remain, including limited non-verbal cues, confidentiality concerns, and perceived difficulties in assessing risk. Remote safeguarding appears to be a context-specific practice shaped by structural conditions rather than a universal solution. Our findings confirm the need to retain an in-person consultation option, and to have clear safeguarding processes and appropriate clinician training. Policies should ensure that remote consultations complement rather than replace in-person care, and further research should examine their impact on safeguarding outcomes and equity, with particular attention to ensuring remote options do not widen existing inequities for digitally excluded populations.

## Data Availability

All data produced in the present study are available upon reasonable request to the authors

## Acknowledgments

We are grateful to all participants who generously gave their time to take part in this study. We also extend our thanks to the members of the study’s Patient and Public Involvement group, the CONNECT Voices panel, for their valuable guidance, including reviewing the interview topic guide and contributing to the interpretation of the findings. We would also like to thank the NHS staff at the three case study sites for their participation in the interviews and for their assistance in facilitating the research.

## References

1. Gibbs J, Solomon D, Jackson L, Mullick S, Burns F, Shahmanesh M. Measuring and evaluating sexual health in the era of digital health: challenges and opportunities. Sexual Health. 2022;19:336–45.

2. Day S, Kinsella R, Jones S, Tittle V, Suchak T, Forbes K. Safeguarding outcomes of 16 and 17-year-old service users of Sexual Health London (SHL.uk), a pan-London online sexual health service. International Journal of STD & AIDS. 2020;31(14):1373–9.

3. Bissessor M, Bradshaw CS, Fairley CK, Chen MY, Chow EP. Provision of HIV test results by telephone is both safe and efficient for men who have sex with men. International Journal of STD & AIDS. 2017;28(1):39–44.

4. Lunt A, Llewellyn C, Bayley J, Nadarzynski T. Sexual healthcare professionals’ views on the rapid provision of remote services at the beginning of COVID-19 pandemic: A mixed-methods study. International Journal of STD & AIDS. 2021;32(12):1138–48.

5. Bosó Pérez R, Reid D, Maxwell KJ, Gibbs J, Dema E, Bonell C, et al. Access to and quality of sexual and reproductive health services in Britain during the early stages of the COVID-19 pandemic: a qualitative interview study of patient experiences. BMJ Sexual & Reproductive Health. 2023;49(1):12–20.

6. McLeod J, Estcourt CS, MacDonald J, Gibbs J, Woode Owusu M, Mapp F, et al. Opening the digital doorway to sexual healthcare: Recommendations from a behaviour change wheel analysis of barriers and facilitators to seeking online sexual health information and support among underserved populations. PLoS One. 2025;20(1):e0315049.

7. NHS England. Safeguarding children, young people and adults at risk in the NHS. 2024. https://www.england.nhs.uk/long-read/safeguarding-children-young-people-and-adults-at-risk-in-the-nhs/#2-introduction-purpose

8. Duffy A, Browne F, Connolly M. Safeguarding adults: A concept analysis. Journal of Advanced Nursing. 2025;81(1):181–97.

9. NHS England. Safeguarding: NHS England; 2023 [updated 26/03/2025. Available from: Available from: https://www.england.nhs.uk/long-read/safeguarding/.

10. Sullivan V, de Sa J, Hamlyn E, Baraitser P. How can we facilitate online disclosure of safeguarding concerns in under 18s to support transition from online to face-to-face care? International Journal of STD & AIDS. 2020;31(6):553–9.

11. Romanis EC, Parsons JA. Early telemedical abortion, safeguarding, and under 18s: a qualitative study with care providers in England and Wales. BMJ Sexual & Reproductive Health. 2023;49(4):238–44.

12. Parsons JA, Romanis EC. ‘All hands on deck’: a qualitative study of safeguarding and the transition to telemedical abortion care in England and Wales. Social Science & Medicine. 2024;348:116835.

13. Bekaert S, Azzopardi L. Safeguarding teenagers in a sexual health service during the COVID-19 pandemic. Sexually Transmitted Infections. 2022;98(3):219–21.

14. Payne R, Clarke A, Swann N, van Dael J, Brenman N, Rosen R, et al. Patient safety in remote primary care encounters: multimethod qualitative study combining Safety I and Safety II analysis. BMJ Quality & Safety. 2024;33(9):573–86.

15. Munro C, Patel R, Brito-Mutunayagam S, Carlin E, Kasliwal A, Manavi K, et al. Standards for Online and Remote Providers of Sexual and Reproductive Health Services. 2019. August 2025 https://www.bashh.org/_userfiles/pages/files/resources/bashhfrshjointstandardsforonlineandremoteproviders2019x.pdf

16. Thornhill J, Bailey A, van Halsema C, Nieman Sims C, Rattue M, Waters L. British HIV Association (BHIVA) guidance for virtual consultations for people with HIV 2021: May 2021. British HIV Association 2021.Accessed: 29/10/2025 Available from: https://bhiva.org/wp-content/uploads/2024/10/BHIVA-guidance-for-virtual-consultations-for-people-with-HIV.pdf

17. Royal College of Obstetricians and Gynaecologists. Best practice in telemedicine for abortion care. London: Royal College of Obstetricians and Gynaecologists; 2022.Accessed: 29/10/2025 Available from: https://www.rcog.org.uk/media/f32nniuk/abortion-care-telemedicine-best-practice-paper-2022.pdf

18. Faculty of Sexual and Reproductive Health, British Association for Sexual Health and HIV. Triage Integration Considerations & Methods to prioritise Vulnerable Groups.; N.D.Accessed: 29/10/2025 Available from: https://www.bashh.org/_userfiles/pages/files/resources/covid/triage_integration_considerations_and_methods_to_prioritise_vulnerable_groups.pdf

19. Spence T, Gibbs J, Wong G, Howarth A, Copas A, Crundwell D, et al. Evaluating the Implementation of Online Postal Self-Sampling for Sexually Transmitted Infections in England: Multisite Qualitative Study. J Med Internet Res. 2025;27:e72812.

20. Palinkas LA, Horwitz SM, Green CA, Wisdom JP, Duan N, Hoagwood K. Purposeful Sampling for Qualitative Data Collection and Analysis in Mixed Method Implementation Research. Administration and Policy in Mental Health and Mental Health Services Research. 2015;42(5):533-44. 21.

21. Witney T, Spurway C, Gibbs J, Munro H, Williams I, Solomon D, et al. The acceptability, appropriateness, and equity of remote consultations in sexual and reproductive health services in England and Wales: a qualitative study. 2026. 10.64898/2026.02.06.26345738.

22. Dixon S, Frost L, Feder G, Ziebland S, Pope C. Challenges of safeguarding via remote consulting during the COVID-19 pandemic: a qualitative interview study. British Journal of General Practice. 2022;72(716):e199–e208.

23. Parsons JA, Romanis EC. The Case for Telemedical Early Medical Abortion in England: Dispelling Adult Safeguarding Concerns. Health Care Analysis. 2022;30(1):73–96.

24. McCarron R, Moore A, Foreman I, Brewis E, Clarke O, Howes A, et al. Remote consultations in community mental health: A qualitative study of clinical teams. Journal of Psychiatric and Mental Health Nursing. 2024;31(5):857–68.

25. Rosen R, Wieringa S, Greenhalgh T, Leone C, Rybczynska-Bunt S, Hughes G, et al. Clinical risk in remote consultations in general practice: findings from in-COVID-19 pandemic qualitative research. BJGP Open. 2022;6(3).

26. Reilly K, Ebersole A. Confidentiality and privacy considerations for adolescents receiving contraceptive health services via telemedicine: A narrative review. Women’s Health. 2024;20:17455057241233126.

27. Wood SM, Pickel J, Phillips AW, Baber K, Chuo J, Maleki P, et al. Acceptability, Feasibility, and Quality of Telehealth for Adolescent Health Care Delivery During the COVID-19 Pandemic: Cross-sectional Study of Patient and Family Experiences. JMIR Pediatr Parent. 2021;4(4):e32708.

28. Humphrey A, Cummins S, May C, Stevenson F. GP remote consultations with marginalised patients and the importance of place during care: a qualitative study of the role of place in GP consultations. BJGP Open. 2025;9(1):BJGPO.2024.0050.

